# Contextual Interaction with Patient and Caregiver Factors for Hospital-at-Home program Engagement

**DOI:** 10.1101/2025.09.24.25336097

**Authors:** Deonni P. Stolldorf, Cathy Maxwell, Amanda S. Mixon, Kemberlee R. Bonnet, Anna L. Sachs, Leah Sommerville, Anna H Gallion, Ashley J. Sellers, Tara B. Horr, Kristina Niehoff, Benjamin French, Eduard Vasilevskis, Catherine H. Ivory, Neesha Choma, David G. Schlundt, Sunil Kripalani

**Affiliations:** Vanderbilt University School of Nursing; Vanderbilt University Medical Center; Vanderbilt University Medical Center, Center for Clinical Quality and Implementation Research; Vanderbilt University Qualitative Research Core; Vanderbilt University Medical Center Home Care Services; Vanderbilt University Medical Center Department of Biostatistics; Vanderbilt University Department of Psychology; University of Wisconsin – Madison, School of Medicine & Public Health, Division of Hospital Medicine; University of Utah College of Nursing; VA Quality Scholars Program, VA Tennessee Valley Healthcare System

**Keywords:** Hospital at Home, context, patient perceptions, implementation

## Abstract

**Background:** Hospital at Home (HaH) is an alternative model of care, where patients who would otherwise be hospitalized are transferred home with hospital care to be carried out in the home environment. Caregivers are central to HaH programs as many patient care tasks shift from hospital personnel to in-home informal caregivers.

**Objective:** To describe the experiences of patients and caregivers, identify factors affecting care delivery, and identify potential patient and caregiver factors serving as barriers and facilitators to engage in a HaH program.

**Study Setting and Design:** **The study was conducted in a large**, academic center in the Southeastern US.

**Data Sources and Analytic Sample:** This qualitative study enrolled a purposive sample of English-speaking patients who received treatment at home and with caregivers of patients who had received HaH services. The Qualitative Research Core conducted 30 to 60 minutes interviews using semi-structured interview guides for patients and caregivers. A total of twenty-three interviews were conducted.

**Principal Findings:** A conceptual framework was developed indicating an interaction between patient/caregiver-level factors (e.g., program attitudes and beliefs) with factors in the broader HaH program context (e.g., provider experience). Modifying factors (e.g., past experiences with hospitalization) affected patient and caregivers’ willingness to engage in the HaH program.

**Conclusions:** Patients and caregivers are willing to engage in HaH but their willingness is affected by factors in the broader caregiving context and person-level factors. Their experience with previous hospitalizations and the COVID-19 pandemic affected their willingness to engage.

## INTRODUCTION

Hospital at Home (HaH) is an alternative model of care, substituting brick-and-mortar care with hospital-level care in the patient’s home. ^1^ In HaH programs, patients are visited daily by members of a multidisciplinary care team, augmented by telehealth technologies between visits to facilitate patient communication.. ^2^ With the HaH approach, many patient monitoring and care tasks (e.g., medication administration) during the day shift from hospital personnel to in-home informal caregivers (hereafter called HaH caregivers), defined as individuals who voluntarily care for family or friends. ^3^

In recent years, the HaH program adoption was accelerated by the Centers for Medicare and Medicaid Services (CMS) waiver and the need of hospitals to address ICU and hospital capacity issues created by the COVID-19 pandemic. ^4,5^ More than 300 hospitals representing 133 systems in 37 states have been approved for HaH programs. ^6-8^ Randomized trials and observational studies have demonstrated improved outcomes – lower complications, length of stay, readmissions, mortality, and cost; and improved functional status, care continuity, and patient satisfaction. ^9-17^

Despite the initial success of HaH programs, many eligible patients choose not to engage in HaH, thus limiting the scope of these programs. The primary barrier to selection include patients’ concern that home-based care would not be sufficient to address their needs while patient perception of their home being unsafe or not therapeutic and concern over the burden of care placed on family members are also reported. ^18,19^ Although some work has been done to understand patient and caregiver perceptions of HaH programs and engagement in associated care tasks, this remains an understudied area. Understanding patient and caregiver perceptions of HaH programs would help healthcare systems and their HaH teams better engage and empower patients and their caregivers in HaH participation.

## STUDY OBJECTIVE

The study objective was to describe the experiences of patients and caregivers to identify factors affecting care delivery and patient and caregiver’s decision to engage in a HaH program. Engagement reflects patients’ willingness to enroll and their satisfaction with the HaH program after enrollment.

## METHODS

### Study Setting, Design and Sample

The study was conducted at a large, academic hospital in the southeastern United States using an observational qualitative design study and a purposive sample. The sample size was based on the number anticipated to achieve data saturation. Patients and caregivers of patients were eligible if they participated in either of the two programs that provided home-based care: the COVID-to-Home (C2H) program or a broader HaH program (see **Table 1)**. As the overall population of HaH patients (and thereby also HaH caregivers) were small due to the program still in the early stages and the limited daily patient census of the program, to enhance recruitment, we included patients and caregivers of hospitalized patients who met eligibility criteria but remained in the acute care setting.

**Table 1:**
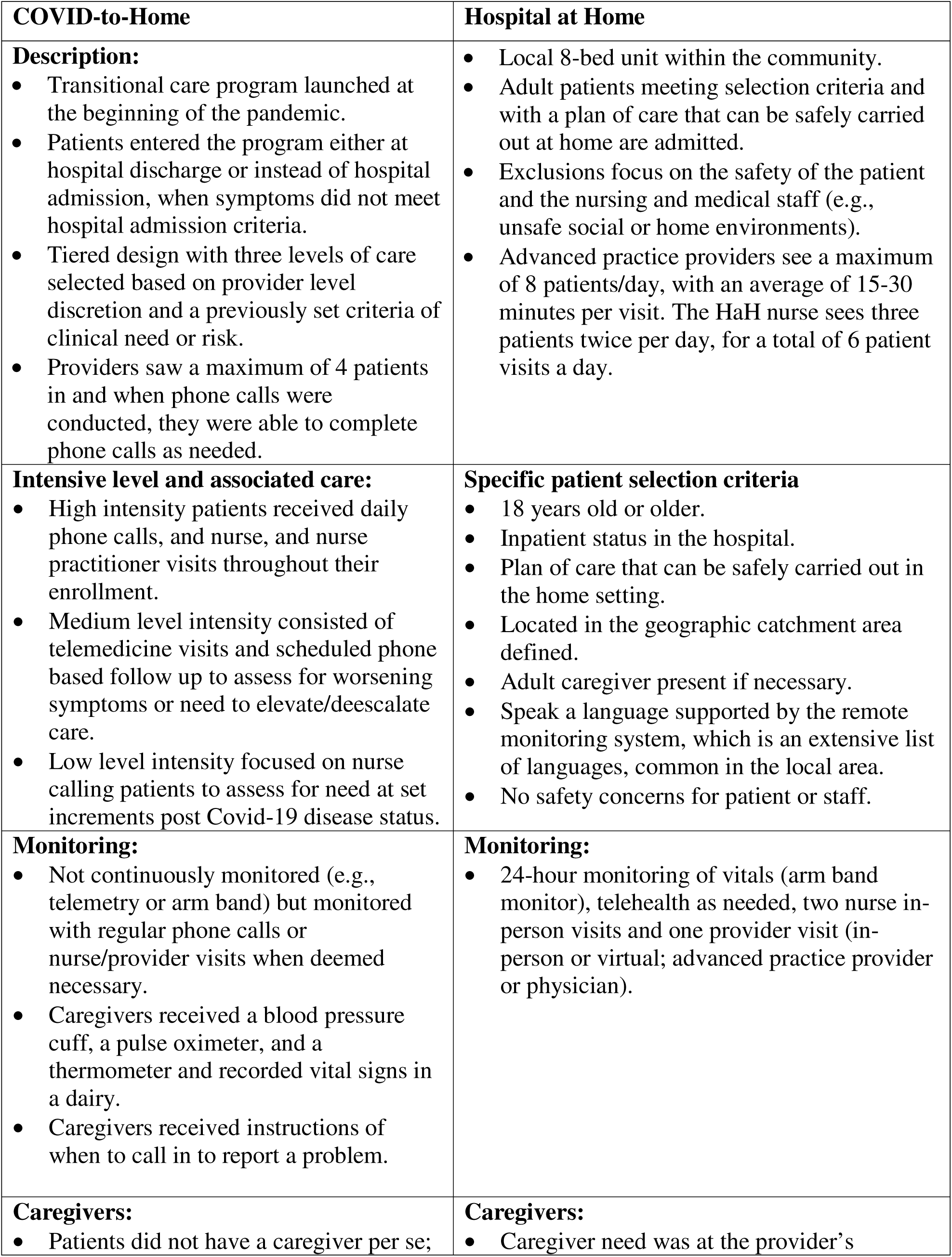
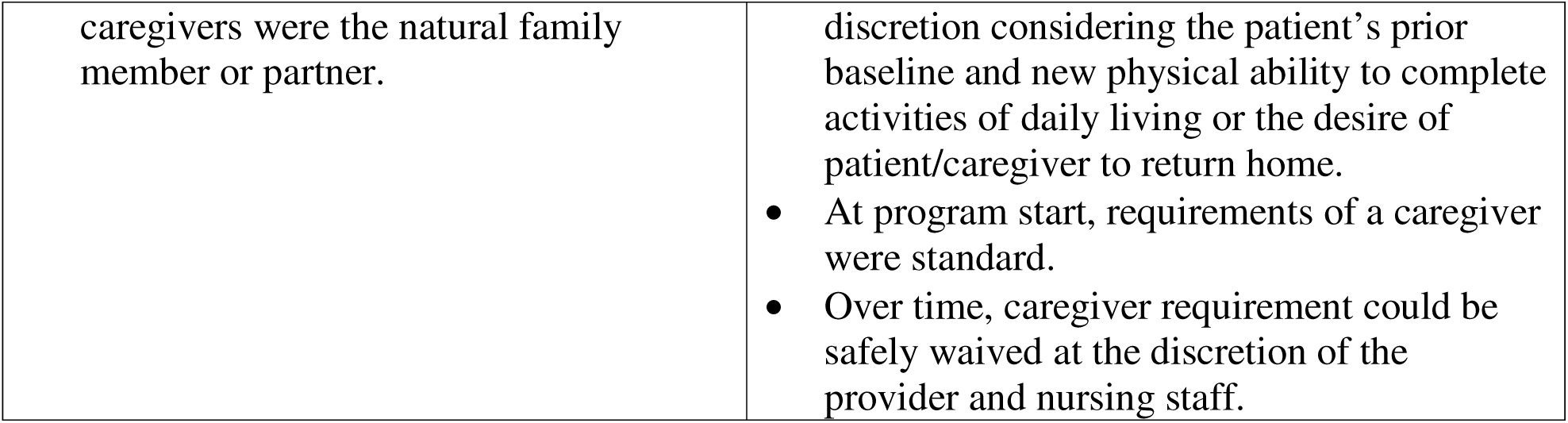
COVID-to-Home and Hospital-at-Home Program.

| COVID-to-Home | Hospital at Home |
| --- | --- |
| **Description:**<br>• Transitional care program launched at the beginning of the pandemic.<br>• Patients entered the program either at hospital discharge or instead of hospital admission, when symptoms did not meet hospital admission criteria.<br>• Tiered design with three levels of care selected based on provider level discretion and a previously set criteria of clinical need or risk.<br>• Providers saw a maximum of 4 patients in and when phone calls were conducted, they were able to complete phone calls as needed. | • Local 8-bed unit within the community.<br>• Adult patients meeting selection criteria and with a plan of care that can be safely carried out at home are admitted.<br>• Exclusions focus on the safety of the patient and the nursing and medical staff (e.g., unsafe social or home environments).<br>• Advanced practice providers see a maximum of 8 patients/day, with an average of 15-30 minutes per visit. The HaH nurse sees three patients twice per day, for a total of 6 patient visits a day. |
| **Intensive level and associated care:**<br>• High intensity patients received daily phone calls, and nurse, and nurse practitioner visits throughout their enrollment.<br>• Medium level intensity consisted of telemedicine visits and scheduled phone based follow up to assess for worsening symptoms or need to elevate/deescalate care.<br>• Low level intensity focused on nurse calling patients to assess for need at set increments post Covid-19 disease status. | **Specific patient selection criteria**<br>• 18 years old or older.<br>• Inpatient status in the hospital.<br>• Plan of care that can be safely carried out in the home setting.<br>• Located in the geographic catchment area defined.<br>• Adult caregiver present if necessary.<br>• Speak a language supported by the remote monitoring system, which is an extensive list of languages, common in the local area.<br>• No safety concerns for patient or staff. |
| **Monitoring:**<br>• Not continuously monitored (e.g., telemetry or arm band) but monitored with regular phone calls or nurse/provider visits when deemed necessary.<br>• Caregivers received a blood pressure cuff, a pulse oximeter, and a thermometer and recorded vital signs in a diary.<br>• Caregivers received instructions of when to call in to report a problem. | **Monitoring:**<br>• 24-hour monitoring of vitals (arm band monitor), telehealth as needed, two nurse in-person visits and one provider visit (in-person or virtual; advanced practice provider or physician). |
| **Caregivers:**<br>• Patients did not have a caregiver per se; | **Caregivers:**<br>• Caregiver need was at the provider's |
| caregivers were the natural family member or partner. | discretion considering the patient's prior baseline and new physical ability to complete activities of daily living or the desire of patient/caregiver to return home.<br>• At program start, requirements of a caregiver were standard.<br>• Over time, caregiver requirement could be safely waived at the discretion of the provider and nursing staff. |

Patients and caregivers were recruited in collaboration with leaders and health care professionals [healthcare provider (HCPs); nurses, advanced practice nurses (APRNs), physicians, social workers, pharmacists) affiliated with the study hospital’s C2H program and/or working on inpatient units at the hospital. Additional recruitment strategies included a Research Notification distribution list, Research Match, Twilio text messaging through REDCap, and in-person recruitment on in-patient units.

### Ethical Considerations

Institutional Review Board (IRB) approval for exempt status was obtained (IRB# 211341) from the local university IRB.

### Data Collection and Analysis

The Principal Investigator collaborated with the local University Qualitative Research Core who conducted the interviews, data analysis, and a preliminary report of the study findings. Phone interviews were 30 – 60-minute duration and guided by semi-structured interview guides for patients and caregivers respectively with questions related to specific domains (see Table 2). Interviews were recorded and transcribed prior to analysis. Participants were compensated for their time.

**Table 2:** Patient and Caregiver Interview Question Domains.

| **Patient Interview Domains** | **Caregiver Interview Domains** |
| --- | --- |
| • Treatment experiences<br>• How they were selected for the program<br>• Medical care received since program completion<br>• Aspects of home or hospital treatment that made recovery easier<br>• Difficulties or challenges<br>• Impact of treatment on quality of life<br>• Caregiver impacts<br>• Experiences with providers, medication, and equipment<br>• Needs and suggestions | • Experiences caring for a family member<br>• Continuation of care after discharge<br>• Supports needed for patients after discharge<br>• Support needed for caregivers specifically; 5) aspects that made caregiving easier; 6) difficulties or challenges; 7) impact of caregiving on quality of life; 8) information needs; 9) experiences with providers, medication, and equipment, and 10) needs and suggestions. |

A hierarchical coding system was developed and refined using the interview guide and a preliminary review of the transcripts and used for both patient and caregiver interviews (see **Table 3)**. Experienced qualitative coders first established reliability in using the coding system on one transcript, reconciling any discrepancies, then independently coded the remaining 34 transcripts. An iterative inductive-deductive approach was used to develop the conceptual framework that is deductively informed by theory while inductively integrating details from the coded qualitative data. ^25^ Deductively, the analysis was guided by the Social Cognitive Theory, ^20^ wherein behavior is influenced by the interaction of personal, environmental, and behavioral factors.

**Table 3:**
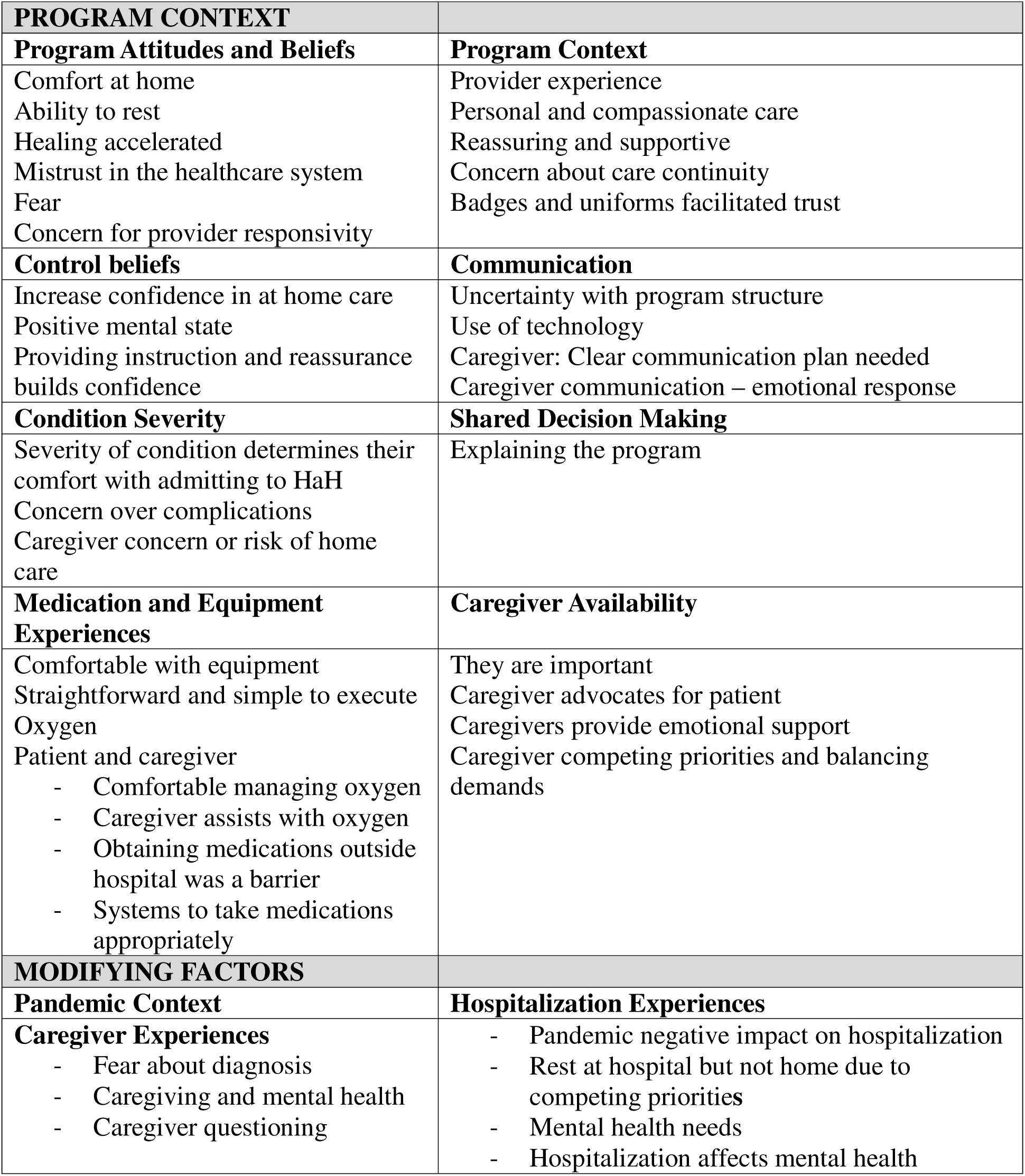
Major Qualitative Categories and Sub-categories.

## RESULTS

### Sample

A total of 35 interviews were conducted, 19 with patients who received treatment at home, either through the COVID-specific or general HaH program and three patients who were eligible for HaH. An additional 10 interviews were conducted with caregivers of patients who had received HaH services and three caregivers of patients who did not participate in HaH.

### The COACHE Conceptual Framework: <u>CO</u>ntextual <u>I</u>nte<u>rAC</u>tion with Patient and <u>C</u>aregiver Factors for Hospital at Home program <u>E</u>ngagement

The conceptual framework developed in this study (**Figure 1)** shows the interaction between person-level and contextual factors affecting patient and caregivers’ willingness to engage in a HaH program. The left displays circles representing patient/caregiver-level factors alongside broader HaH program context. The middle arrows indicate factors that modify willingness to engage. The outcome, “Willingness to Engage,” reflects overall engagement and satisfaction with the HaH experience. For participant quotes related to these factors, see **Table 4**.

**Figure 1:**
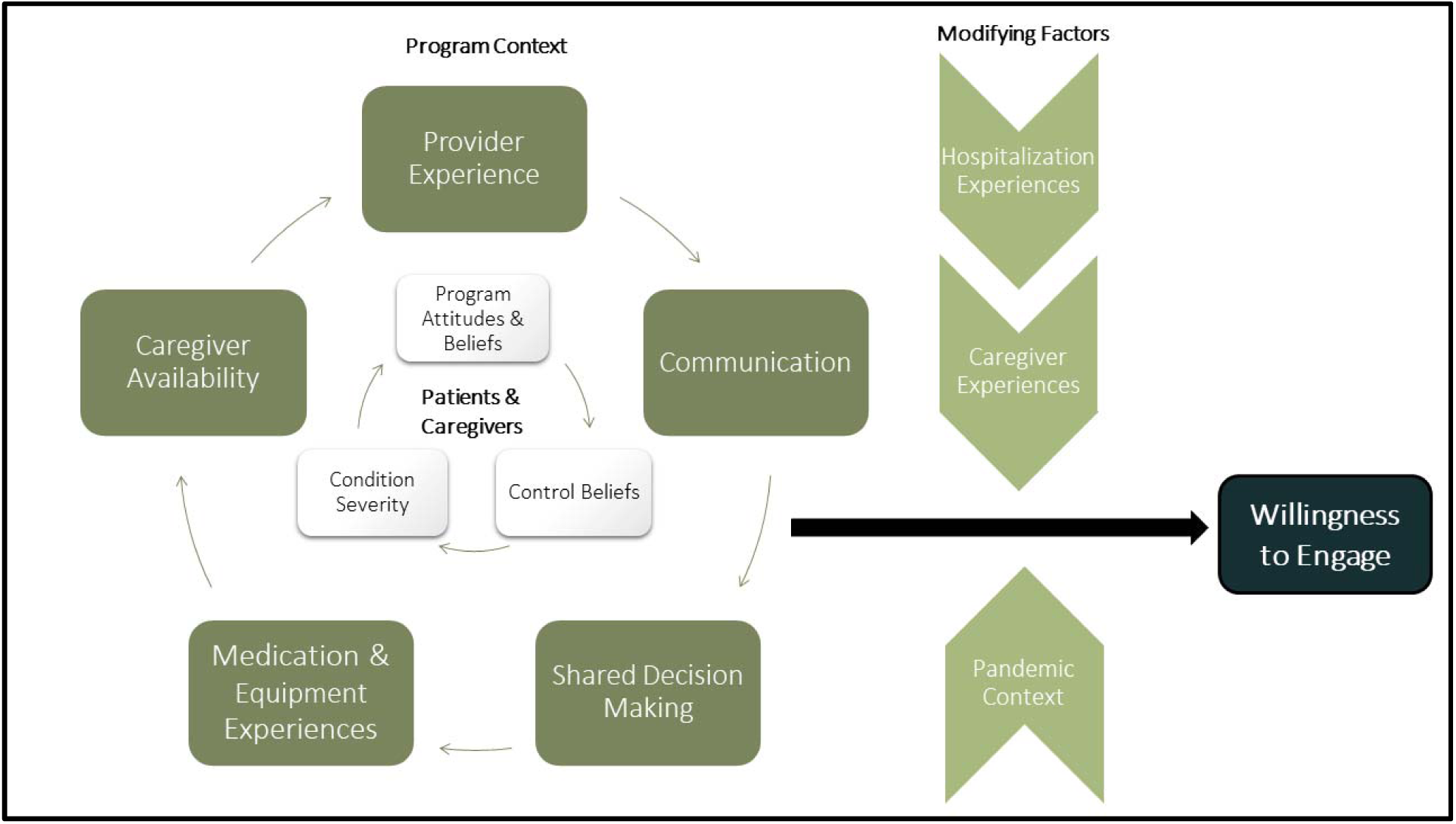
Conceptual Framework for Understanding Willingness to Engage with the HaH Program.

**Table 4:**
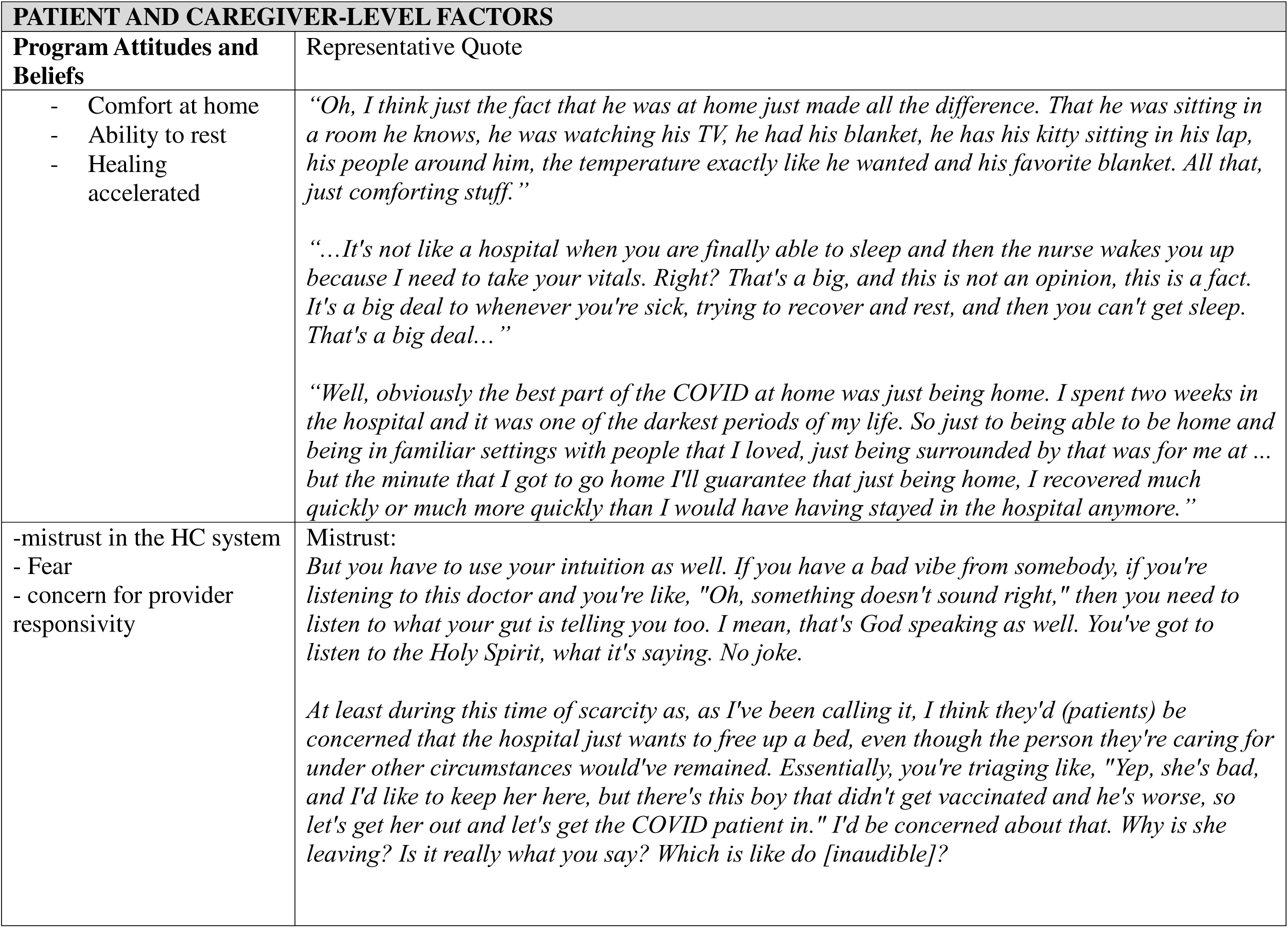
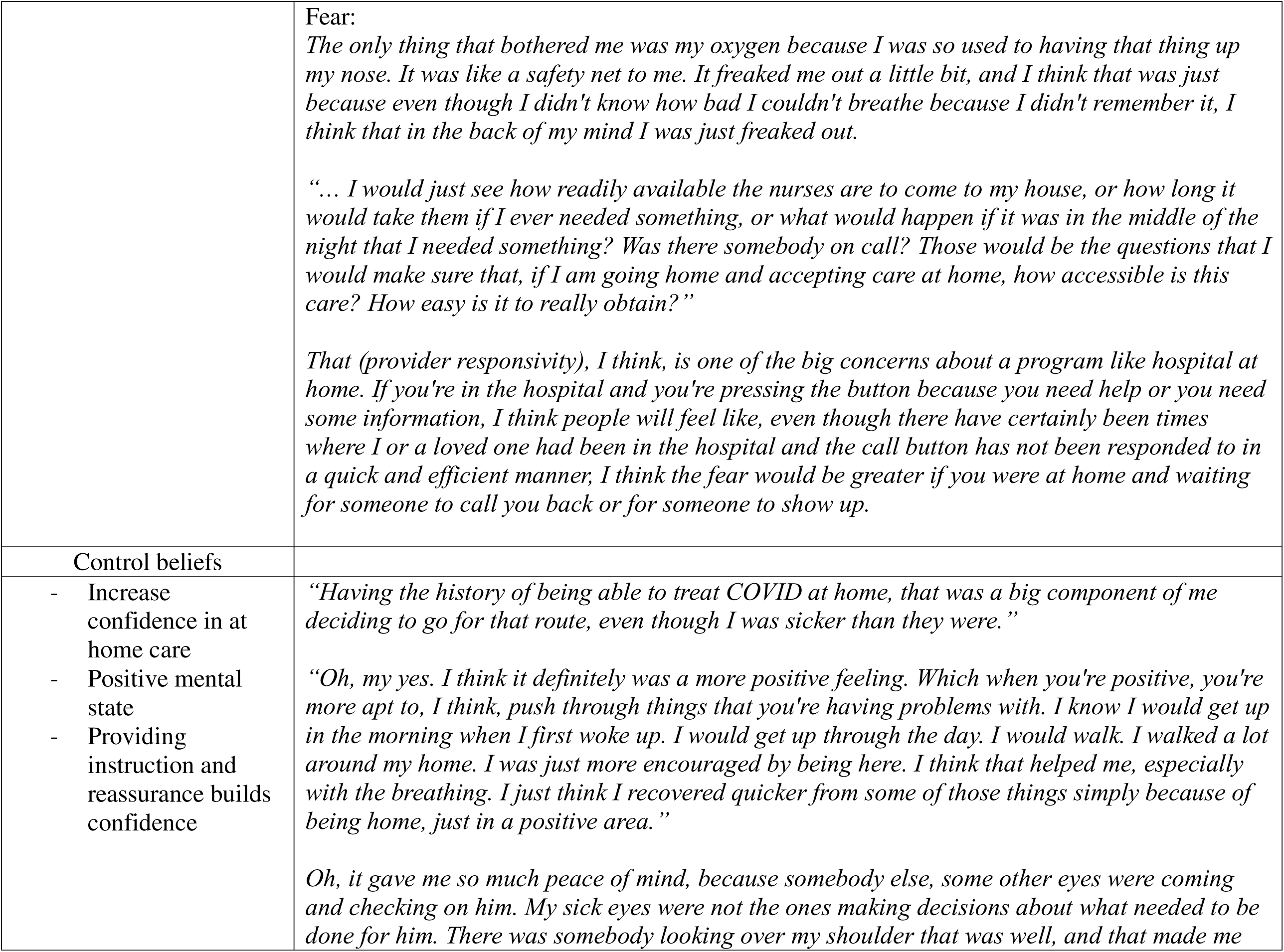
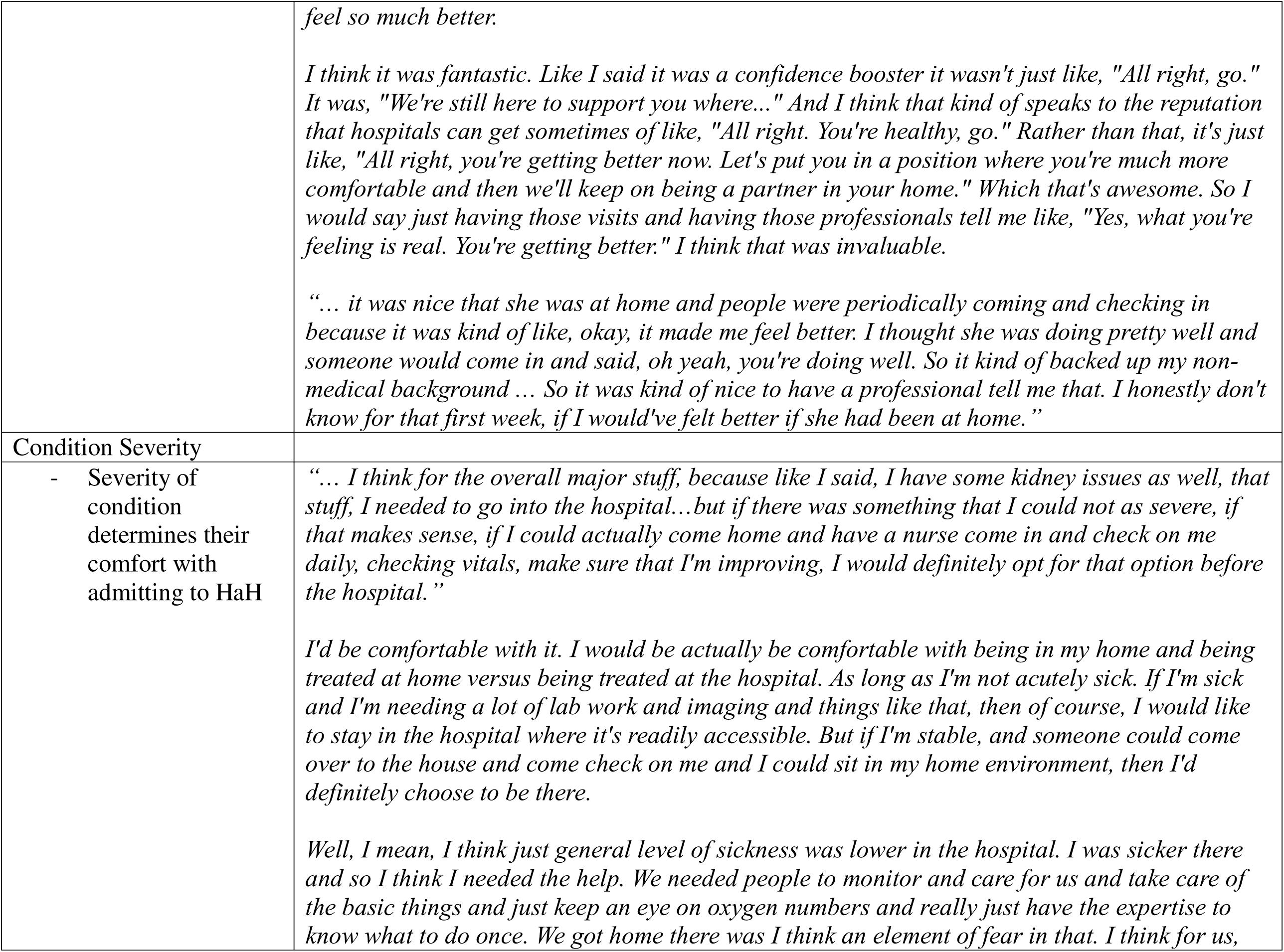
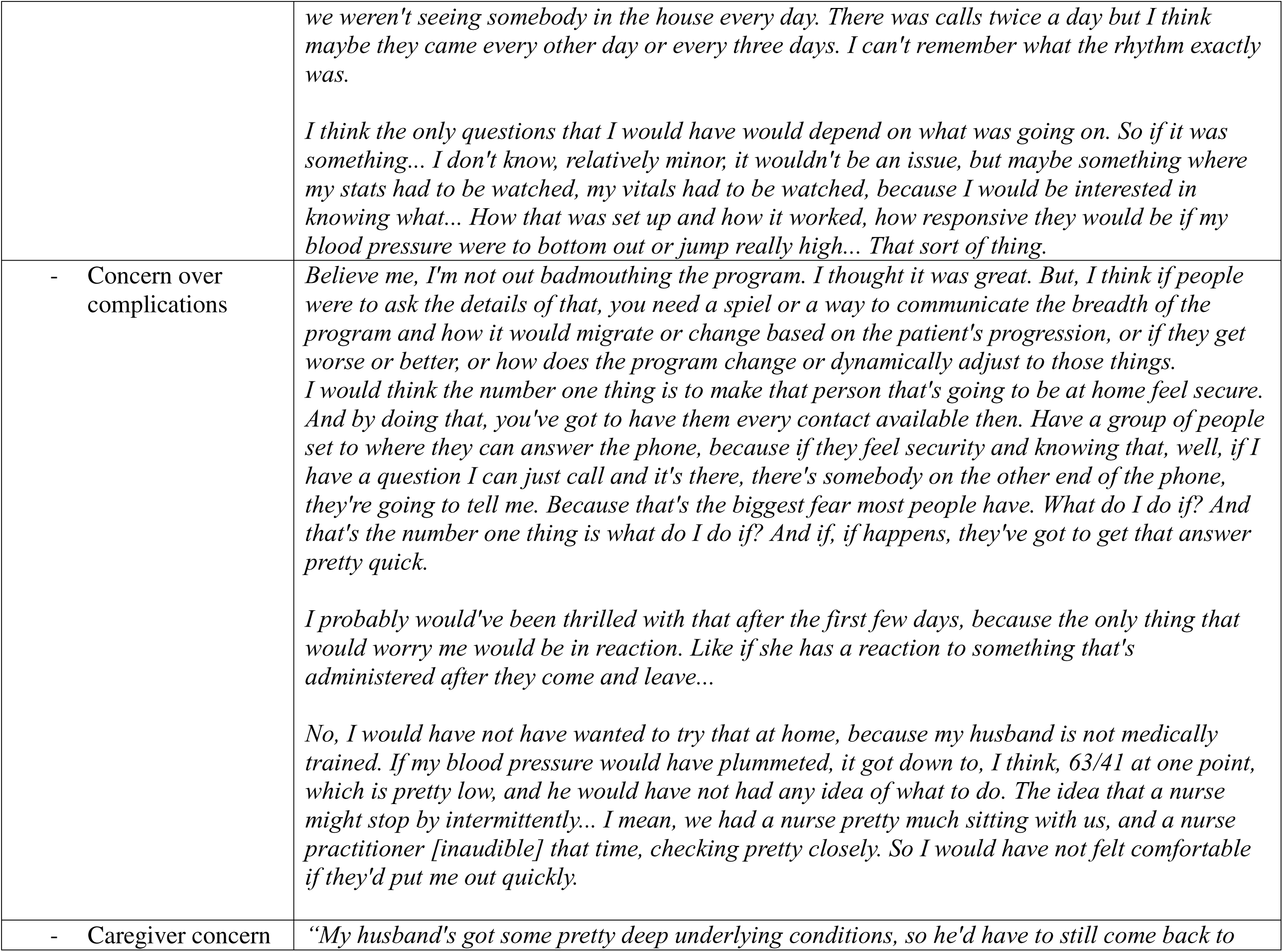
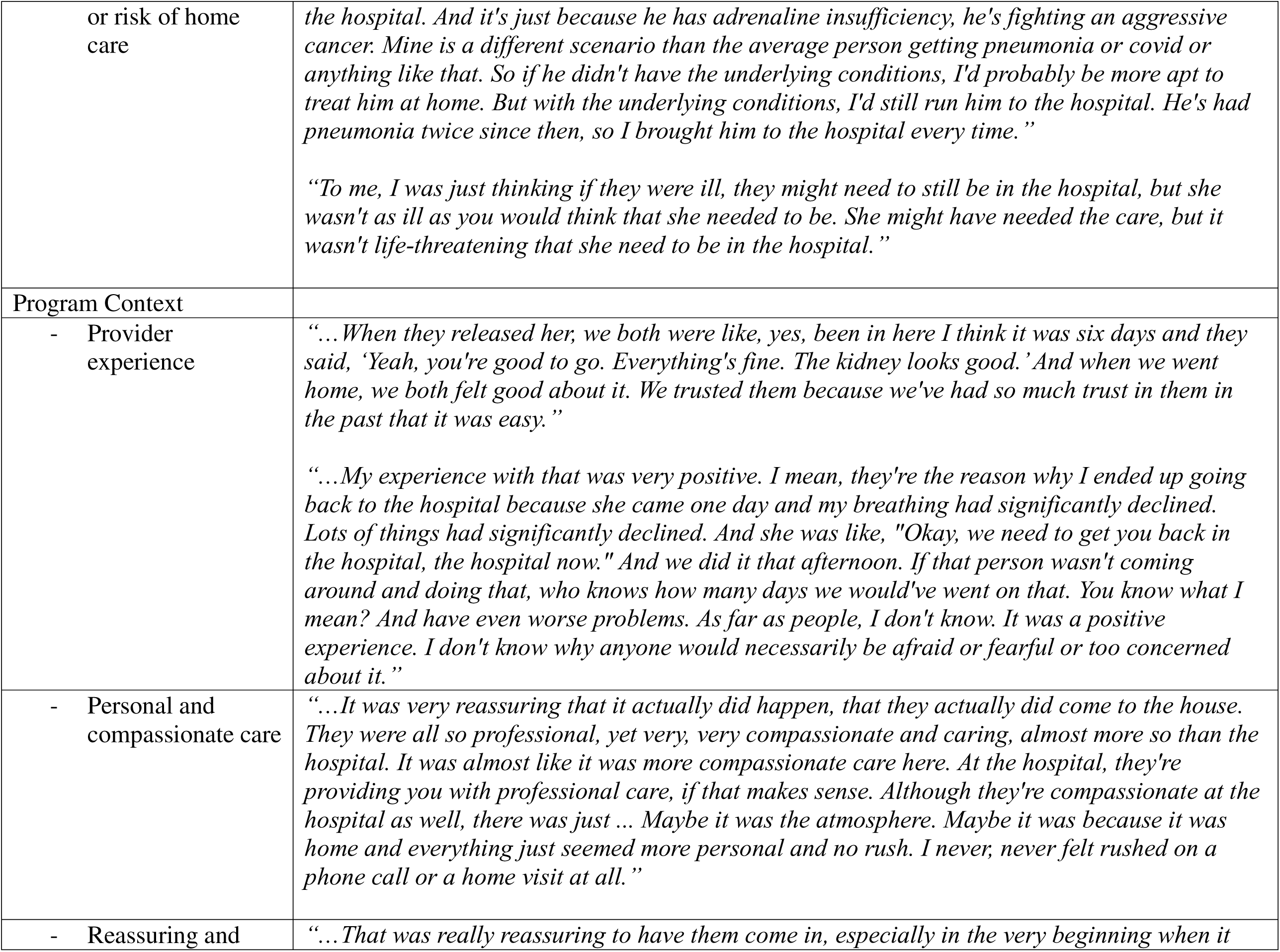
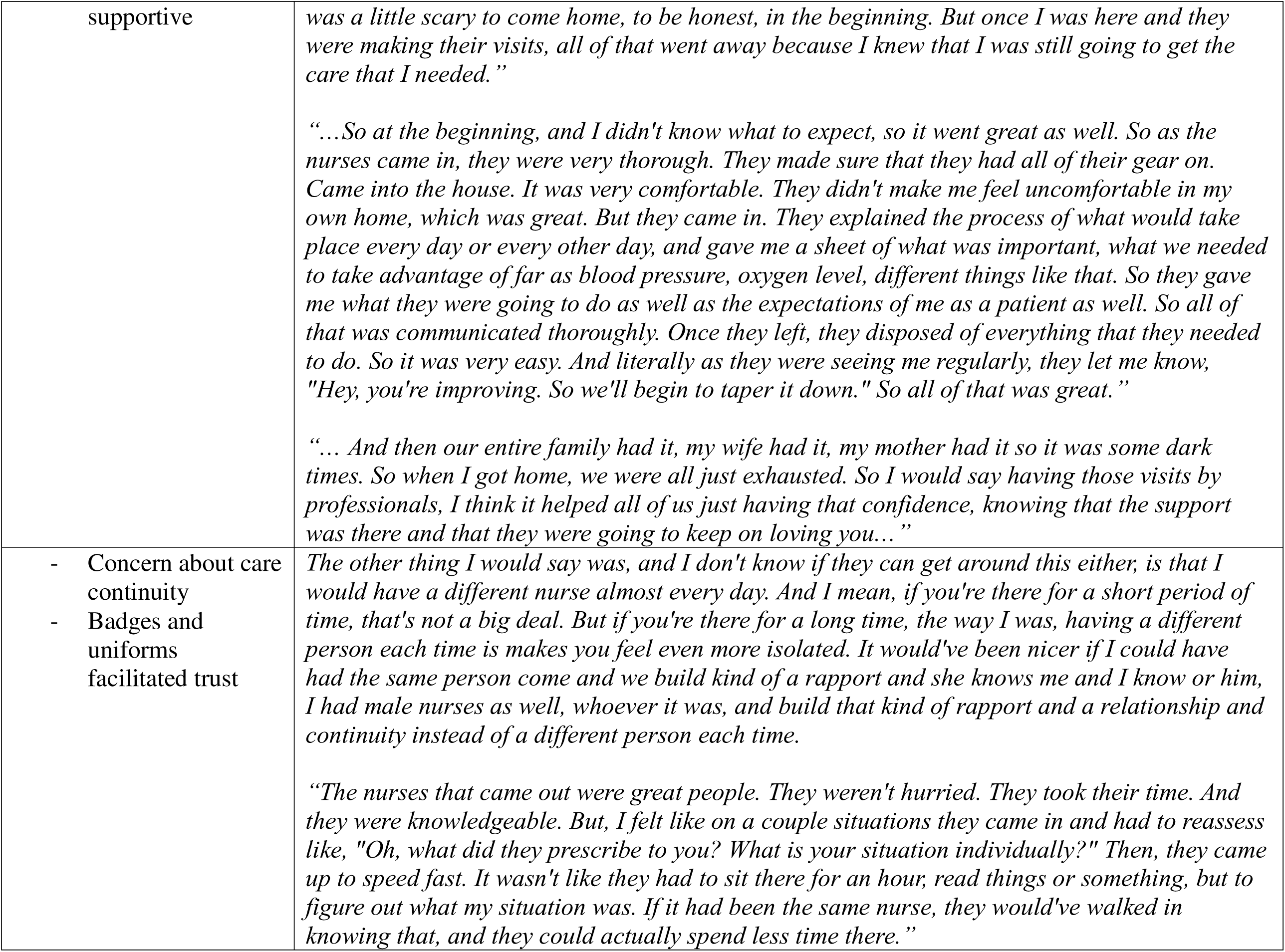
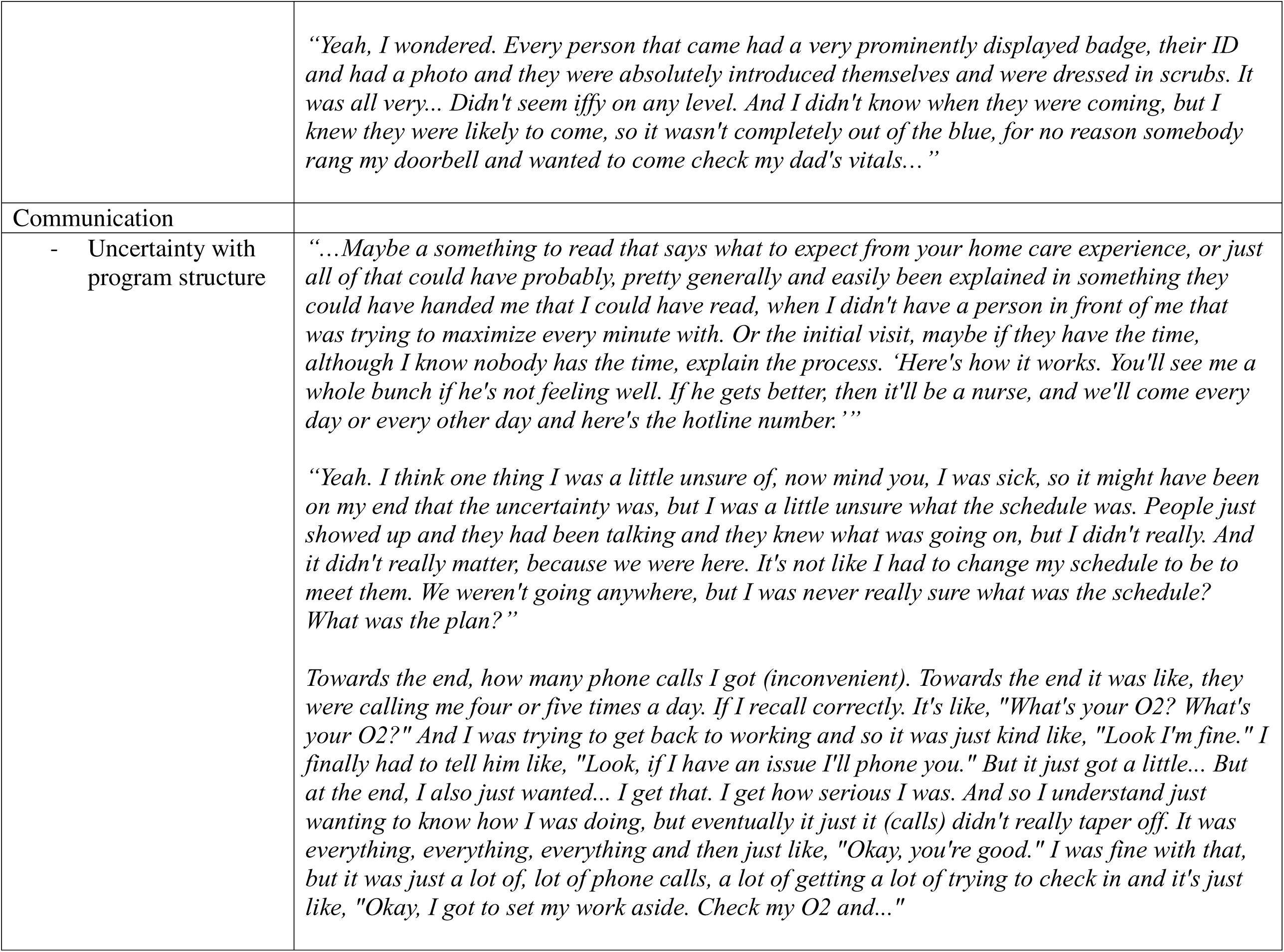
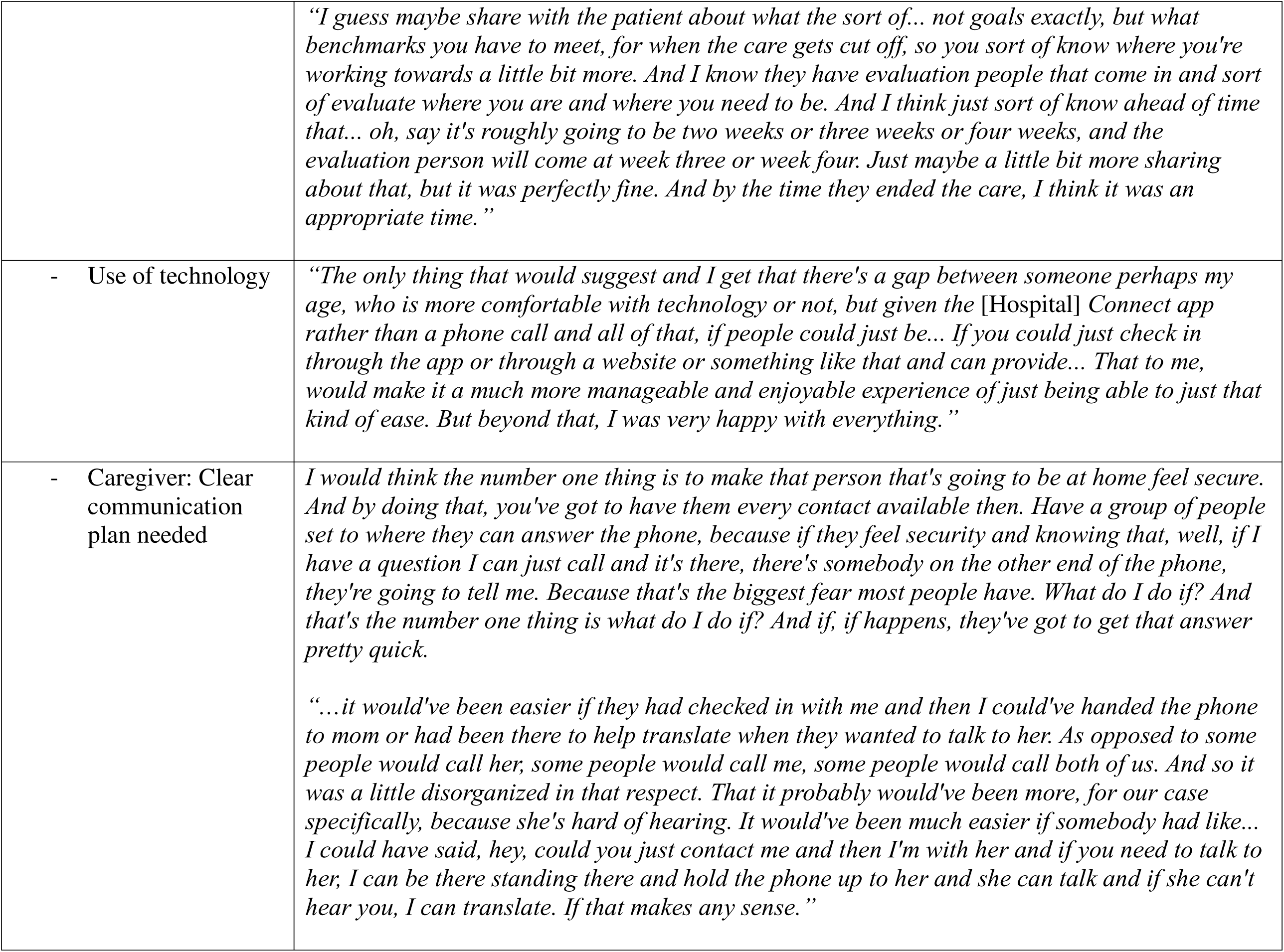
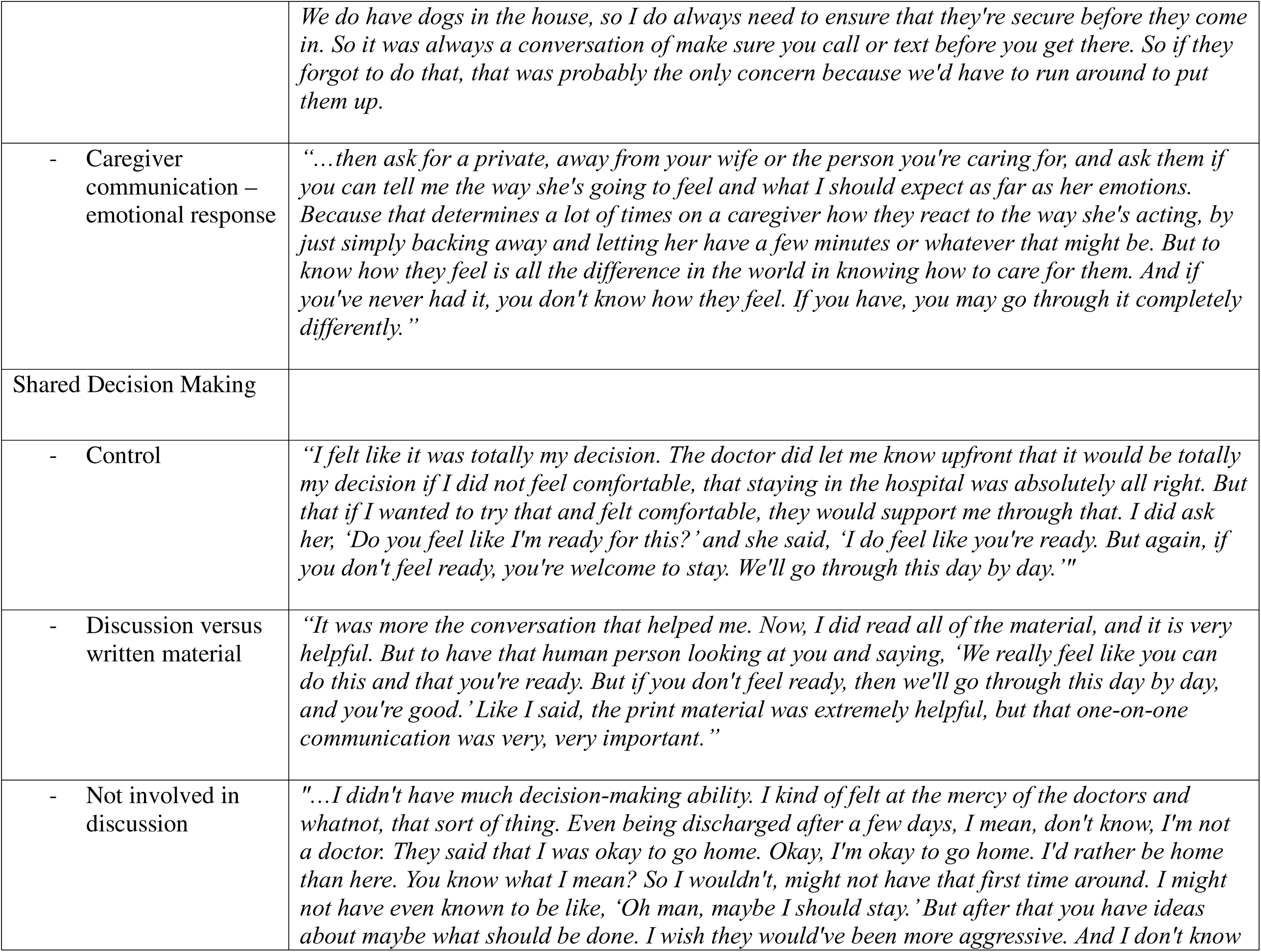
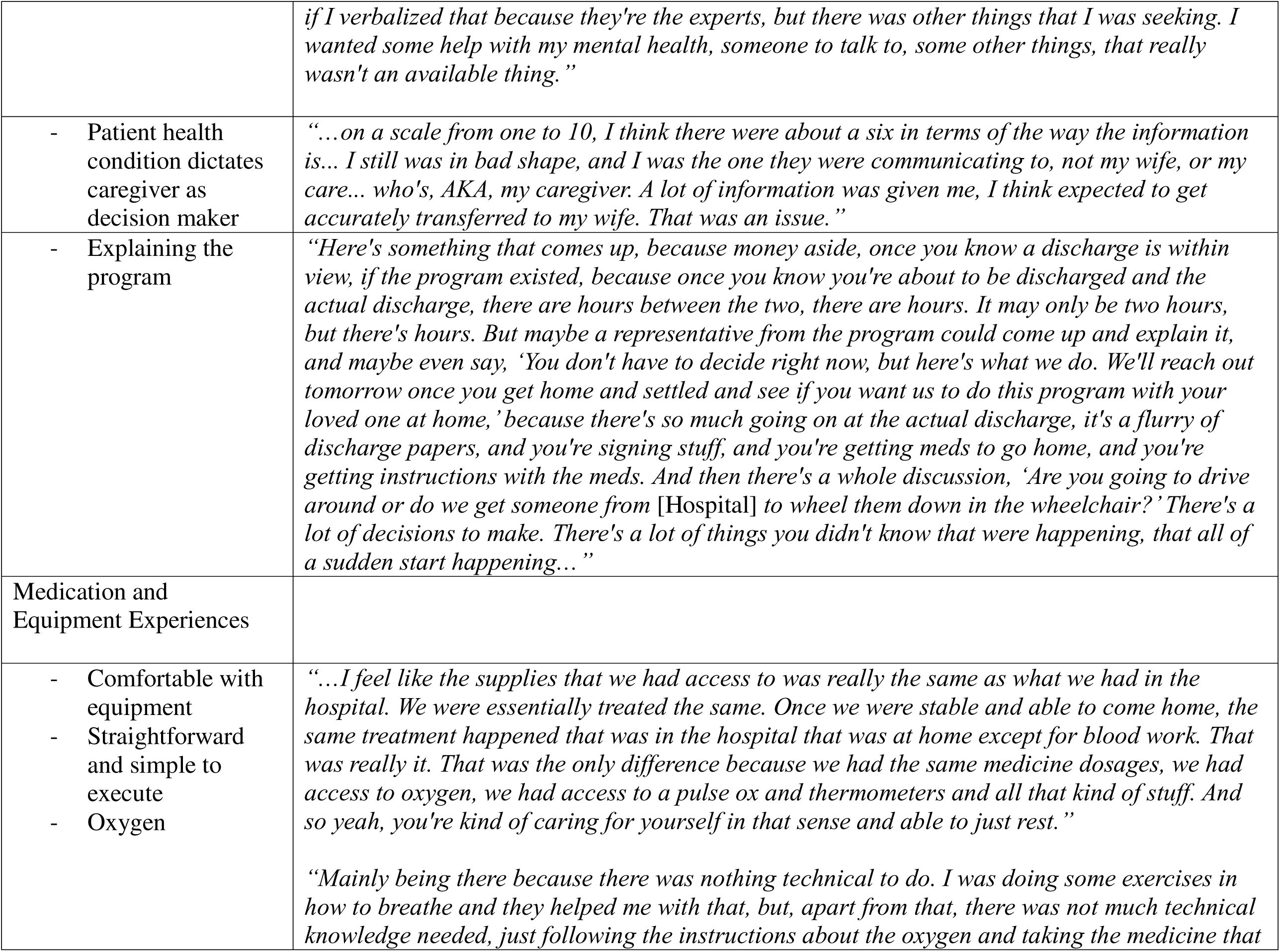
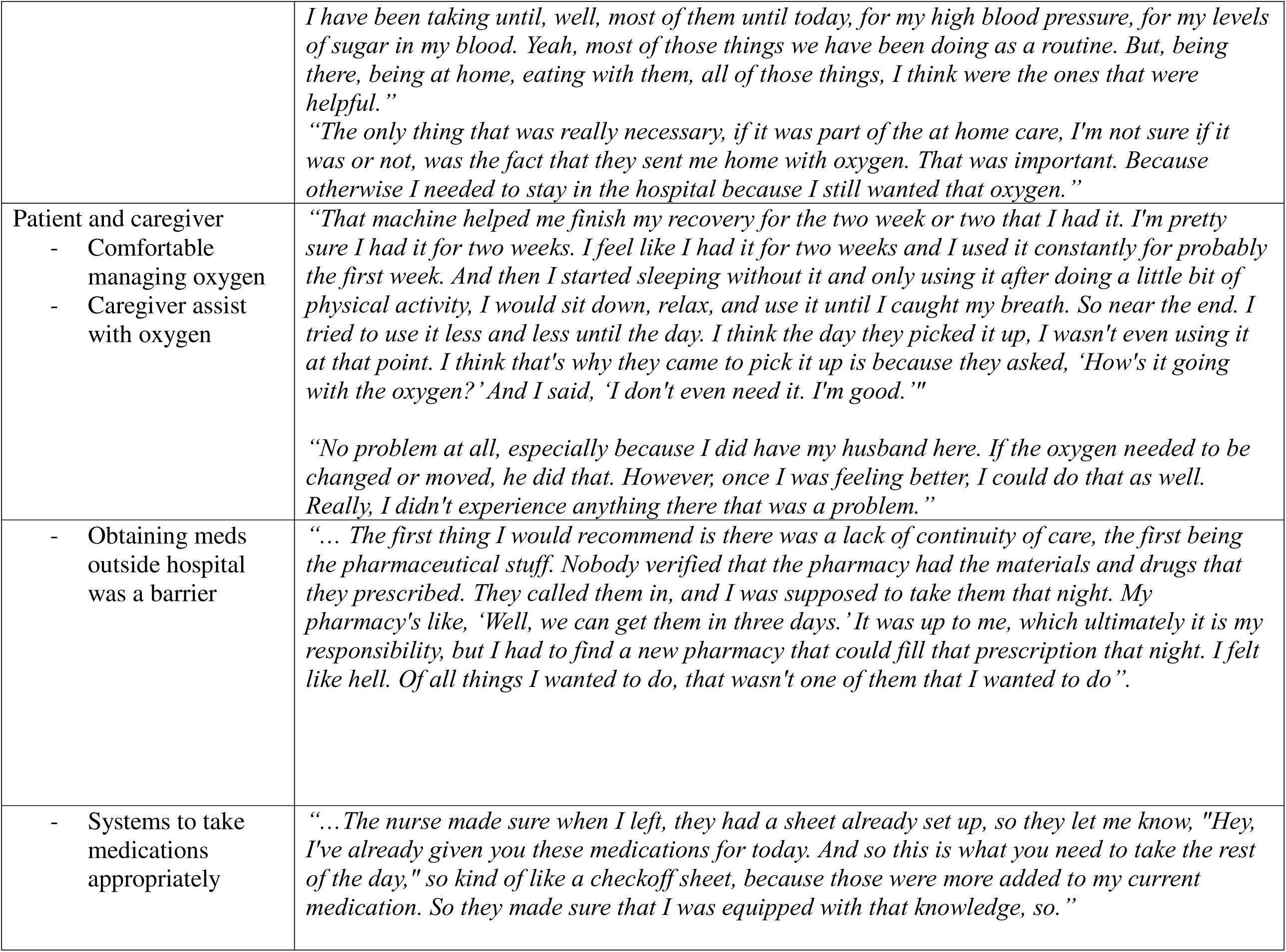
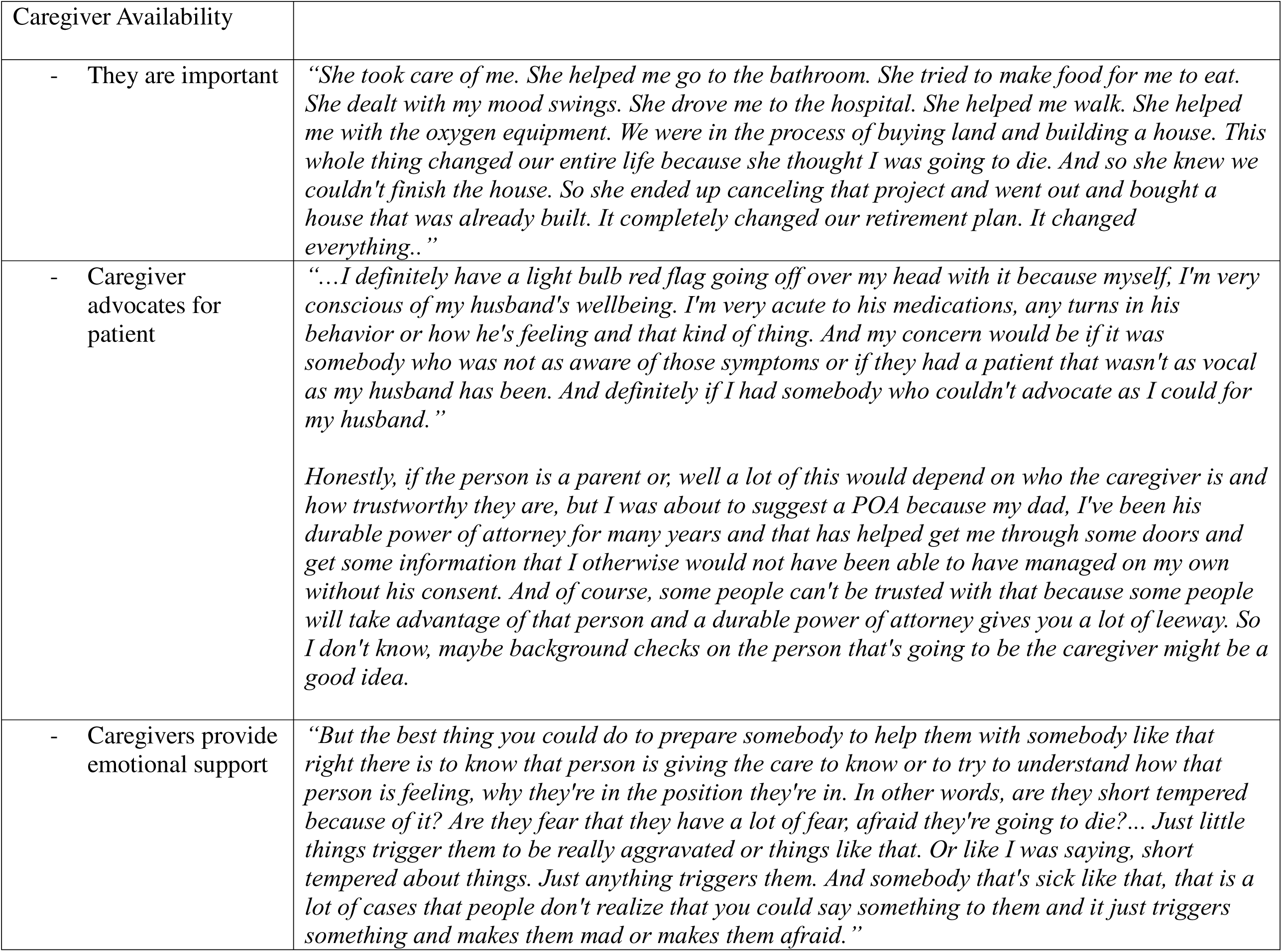
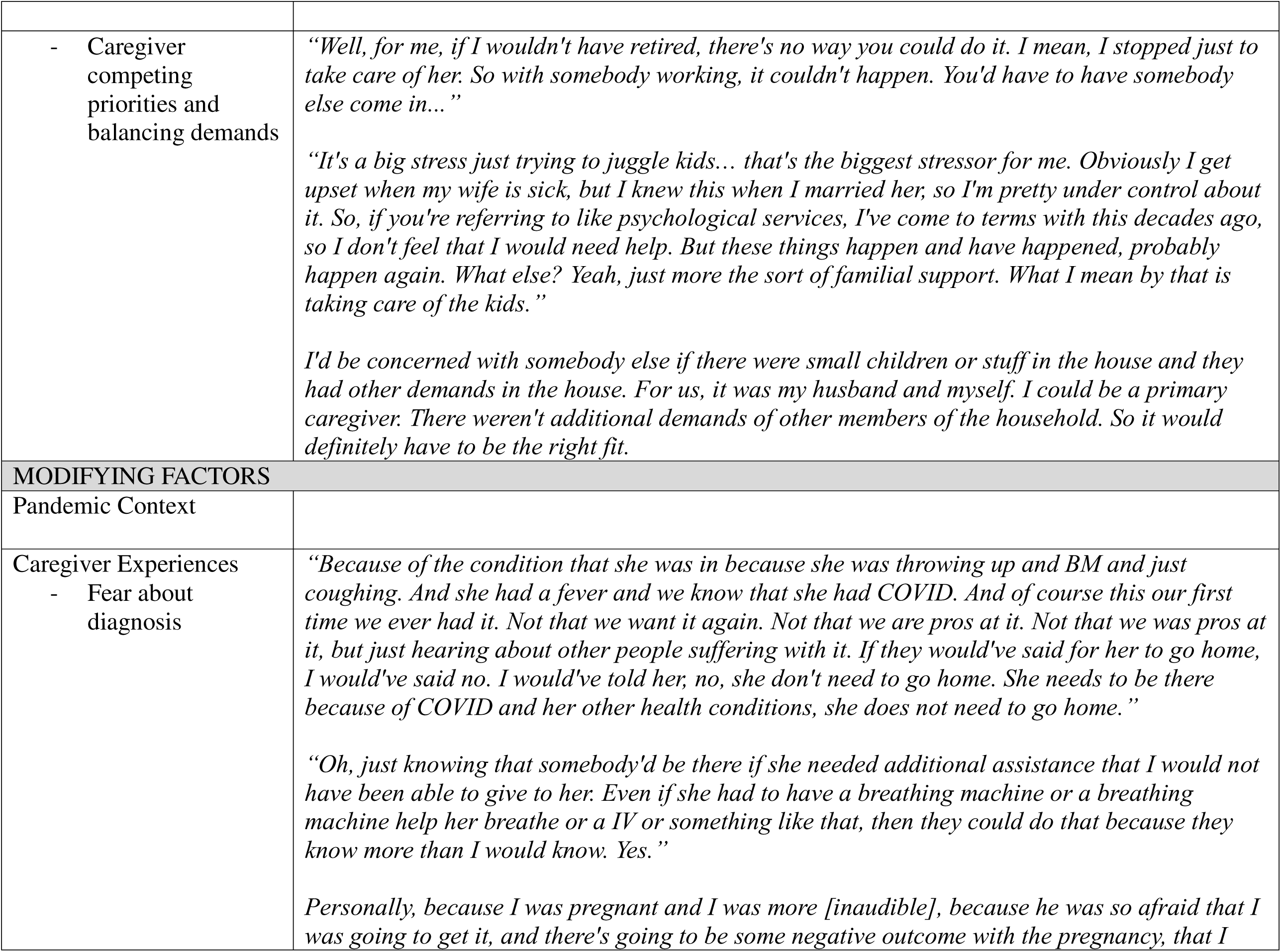
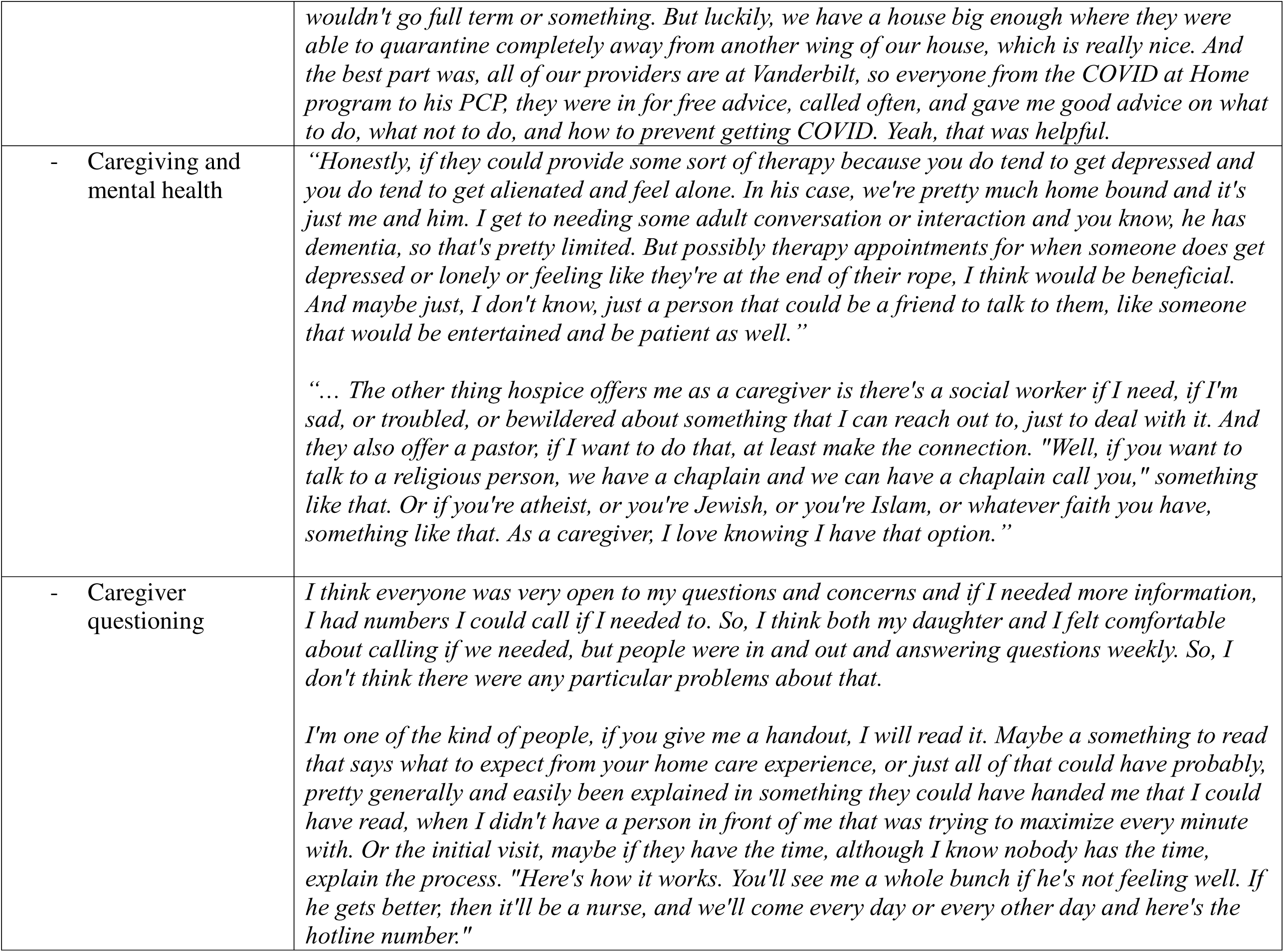
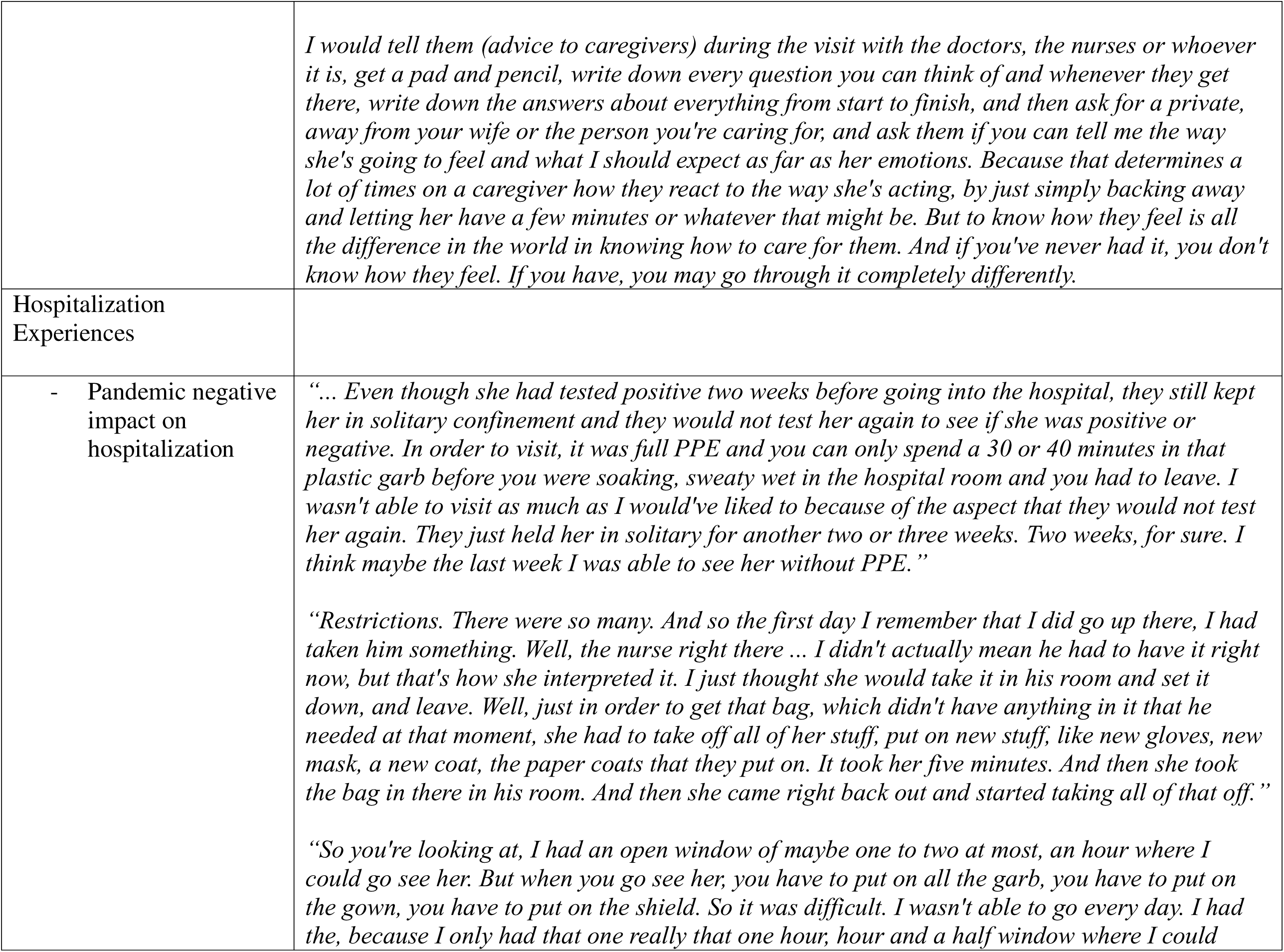
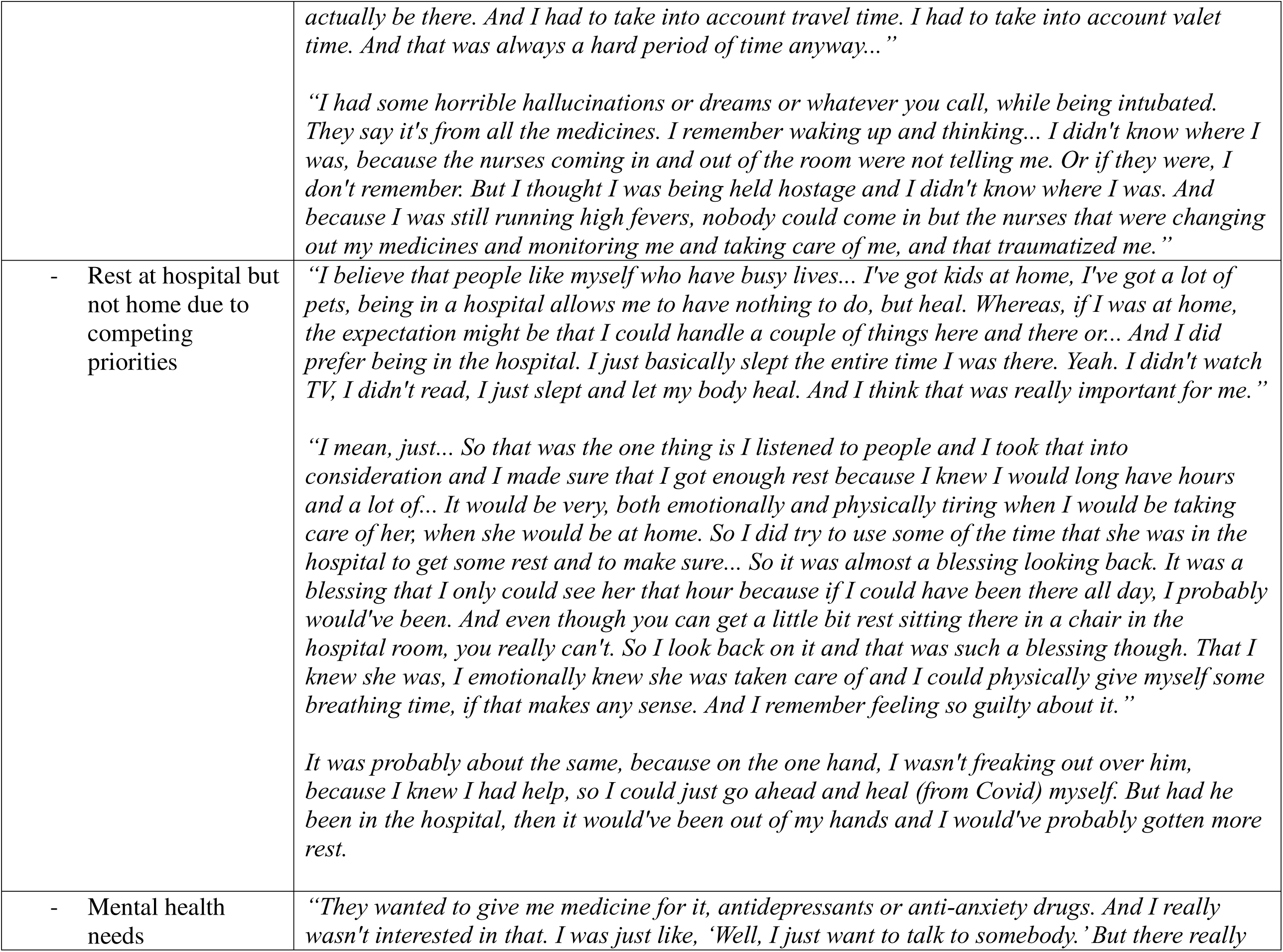
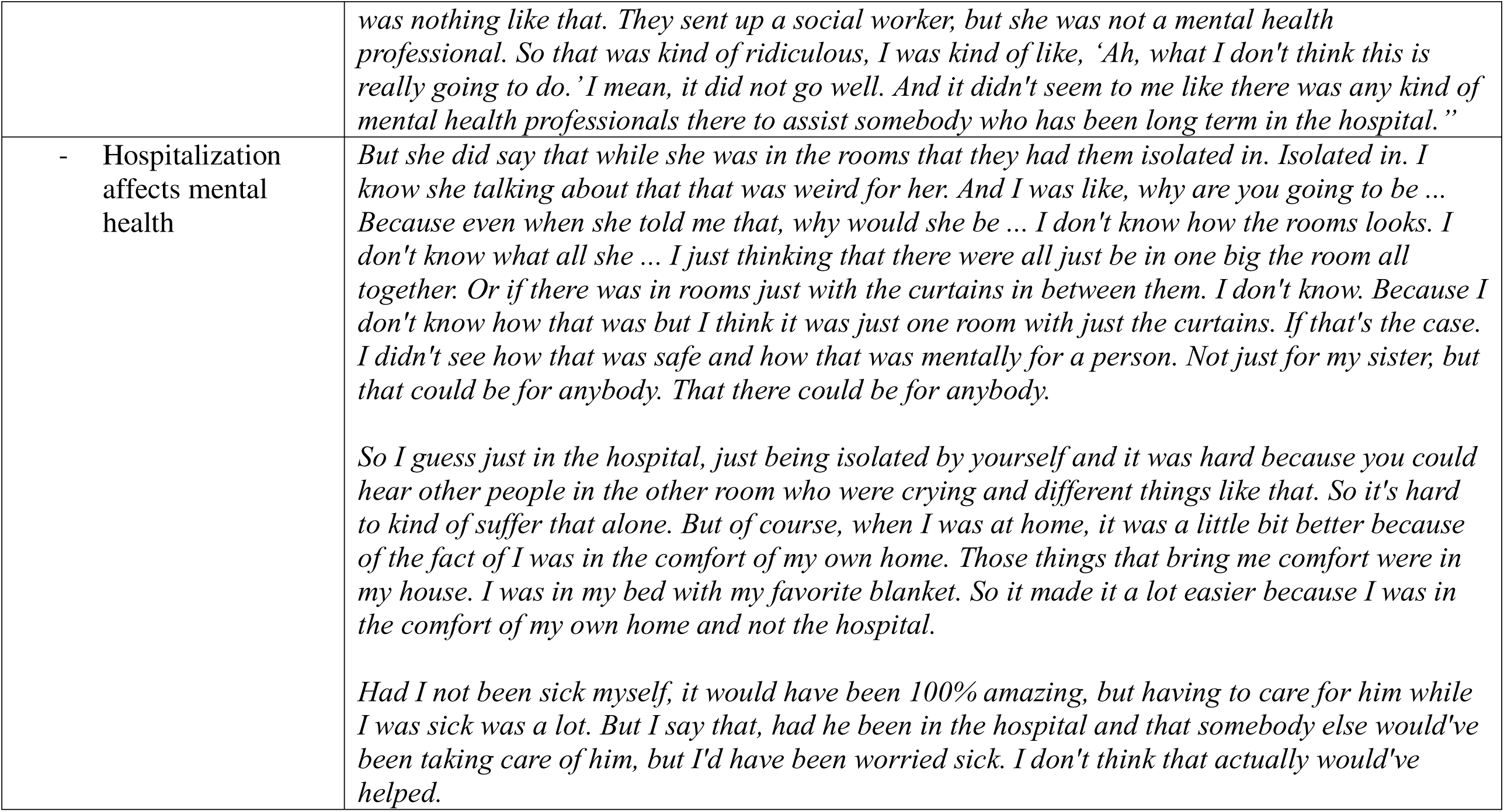
Qualitative themes and representative quotes.

### Patient and Caregiver-level Factors

#### Program Attitudes and Beliefs

Participants expressed varied attitudes and beliefs toward HaH programs. Many felt the “comforts” of home facilitated healing and their mental health, including access to preferred meals and personal items, the ability to receive visitors, and improved rest due to the familiar surroundings and lack of disruptions at night. Some participants, however, expressed feelings of fear of being outside a brick-and-mortar facility and mistrust as to whether HaH programs were truly beneficial for them or for the healthcare system. Several participants expressed fear related to not being continuously monitored at home and concerns regarding provider responsiveness.

#### Control Beliefs

Participants’ confidence in their ability to treat their illness at home and feeling some control over the outcome indicated their willingness to engage in the program. Most patients and caregivers felt confident in following the necessary HaH procedures. Participants noted their improved mental health in the home environment increased their motivation to heal. Providers enhanced participants’ confidence through instruction and reassurance.

#### Condition Severity

Many participants’ support of and active engagement with HaH programs was contingent on the severity of the condition being treated with conditions at low-risk for complications seen as suitable for HaH care. Caregivers expressed concerns about the risks associated with home care for more severe conditions.

### Program Context

#### Provider Experience

Most participants had positive experiences and provided feedback on what went well along with suggestions for improvement. For those transferred to the HaH following hospitalization, a positive provider experience while hospitalized facilitated a willingness to engage in the HaH program. Participants’ experiences with having providers in their home was positive, with care perceived as more personal and compassionate and reassuring and supportive. Most participants expressed feeling comfortable with having different providers every day, though some noted they needed to provide treatment updates for new providers and expressed a preference for having the same providers throughout the program. One caregiver identified that the prominent badges and uniform facilitated trust in the providers. Having well-trained, well-prepared staff is seen as an essential component of HaH care.

#### Communication

Participants described communication as an integral part of the HaH program but noted gaps in communication. They described uncertainty with the program structure and expressed a need for communication regarding program expectations. Most participants felt comfortable with the use of telehealth communication and the use of an application (app) and phone calls. While patients felt that communication was adequate once they were familiar with the program structure, caregivers identified additional communication needs. One caregiver described inconsistencies in who the providers were communicating with, and the need for a clear communication plan. One participant felt it was important for providers to communicate with caregivers about potential emotional responses from the patient.

#### Shared Decision Making

Participants responses with their involvement in the decision to receive hospital care at home varied from active participants in the decision to little to no involvement. Discussion with providers were seen as more helpful than written material. Other participants felt they were not involved in the decision but should have been and disagreed with some decisions that were made. One participant did not feel they were able to engage in the process due to their health condition, and that the decision should have been made with their caregiver instead. Some of the participants felt the busy hospital environment at the time the program was presented to them made it difficult to decide to engage. One caregiver suggested the hospital have a representative to explain the program during downtime.

#### Medication and Equipment Experiences

Hospital at Home programs require various levels of medication and equipment management from patients and caregivers. Most were comfortable with their equipment. Both patients and caregivers were also comfortable with managing the oxygen equipment. Managing medication schedules was also described as straightforward. Distinct to COVID-to-Home patients, they noted that obtaining the medications outside of the hospital system was a common barrier due to either many pharmacies being closed at the time of transferred to HaH or they did not have the needed drugs in stock. This was not a concern for HaH patients as they receive their medication from the HaH program. Participants described systems their providers put into place to ensure drugs were taken appropriately, including pill packs and instruction sheets.

#### Caregiver Availability

Caregivers were identified as an important part of the HaH program. One patient described the importance of their caregiver as someone who took care of her, helped her around the house, prepared foods, and assisted with transportation. Another participant, a caregiver, expressed concern for patients who do not have caregivers available to advocate for them as they perceived advocating for the patient’s needs was important. Participants, particularly caregivers, commented on the importance of emotional support from caregivers. Some caregiver participants stated that their availability was negatively impacted by competing priorities such as work or children.

### Modifying Factors

#### Pandemic Context

The COVID-19 pandemic had various effects on hospital policies, procedures, and emergency room care. Patients and caregivers described requirements for personal protective equipment (PPE), visitation restrictions, and limited availability of beds during surges. The following modifying factors were heavily impacted by this pandemic context.

#### Caregiver Experiences

Many caregivers had experiences caring for their family members long-term. These experiences modified their willingness to engage in HaH program, which influenced the patients’ decision. Caregivers noted the negative effects of caregiving on their mental health and the need for the healthcare system to support this impact. Another caregiver, indicated mental health services provided benefited them while caring for their family member. Caregivers who had not experienced HaH programs had many clarifying questions, and most were curious what support was provided and how communication with providers was established. The COVID pandemic was frightening for caregivers, and involved isolation, loneliness, anxiety, and depression. Support for caregiver distress could be an important element of HaH programs.

#### Hospitalization Experiences

Hospitalization experiences varied among participants, with a spectrum of negative and positive examples described. These experiences modified the patients’ and caregivers’ willingness to receive HaH care. For many, the COVID-19 pandemic made hospitalization a negative experience and facilitated support of the HaH program. Negative hospitalization experiences resulted from the use of protective equipment and limited visiting hours. For some patients and caregivers, hospitalization allowed for more rest, which would not have been possible at home due to competing priorities. The mental toll of hospitalization was also discussed and the lack of much needed mental health support for patients in the COVID to Home (C2H) program. A caregiver noted the negative effect of hospitalization on her mother’s mental health and believed her mother would have been better if she had been treated at home.

## DISCUSSION

This study identified factors affecting patients and caregivers’ willingness to engage in HaH programs, including person-level and contextual elements. Patients and caregivers recognized HaH programs improved patients’ comfort and rest and accelerated their healing and likely contributed to program satisfaction. Their positive attitudes and beliefs align with previous research demonstrating the benefits of receiving care at home. ^12,21^ A survey of 214 patients (HaH:84; acute care hospital:130) indicated high satisfaction with HaH care among patients and family members when compared with inpatient care. ^21^ Additionally, a recent Cochrane review reported similar findings of high patient satisfaction with HaH programs compared to in hospital. ^17^ Previous studies also noted that HaH patients typically experience better sleep, increased physical activity, and more favorable care transitions compared to hospitalized patients.. ^15^ However, similar to previous studies of patient perceptions of the risks and benefits of home care, ^19,22,23^concerns over safety, especially for patients with acute conditions, were seen as a potential barrier to engagement.

Patient and caregiver mistrust the healthcare system and fear of the potential lack of care and provider responsiveness when needed was offset by positive experiences during previous hospitalizations. Mistrust in the healthcare system is associated with failure to take medical advice, keep follow-up appointments, and failure to fill prescriptions. barrier. ^24^ However, providers’ use of patient-centered communication skills and responsiveness to patients’ healthcare needs might buffer the effects of mistrust. ^25^ Therefore, comprehensive patient and caregiver education and transparent, patient-centered communication when HaH admission is discussed is crucial for patient and caregiver engagement in HaH. Increased healthcare system transparency will further foster trust and give providers a better understanding the needs of the communities in which they work. ^24^ Moreover, instruction and reassurance about HaH program components and care delivery will foster patients and caregiver confidence to engage in these programs. Their confidence in managing their illness at home underscored the role of self-efficacy in patient engagement with HaH programs. In turn, high self-efficacy likely increases their resilience to stressful situations while in HaH. ^26^ Condition severity is an important driver for patient and caregiver program acceptance, necessitating transparent and standardized criteria for patient selection to HaH programs. Future studies should identify standardized, evidence-based patient criteria for HaH admissions that enhance patient safety and limit the need for escalating care. Research is also needed to examine robust communication strategies to align patient and caregiver expectations with the capabilities of HaH programs to address patient and caregiver barriers.

The delivery context of HaH significantly influences patient and caregivers’ willingness to engage in the HaH program. Prior positive healthcare experiences enhanced their willingness, further enriched by the personal and compassionate care they received. Effective communication, shared decision making, medication and equipment experiences, as well as caregiver availability are pivotal to their engagement. Our finding of adequate communication enhancing patient and caregiver’s feelings of reassurance, support, and comfort aligns with reports of effective communication strategies enhancing patient and caregiver satisfaction and comfort with HaH programs. ^27^ This might be because active involvement in shared decision-making facilitated patient autonomy and their greater willingness to engage in HaH care.

Caregivers reported the need for clear, consistent provider communication and caregiver involvement with provider-patient communication to foster both patient and caregiver engagement with HaH. ^25^ This finding aligns with a recent study which demonstrated patient and caregivers’ desire for collaborative discharge planning and for healthcare providers to anticipate the patient’s self-care needs. ^28^ Discharge instructions can also serve as a communication medium and should include caregivers. ^29^ Future research studies are needed to identify the best practices for shared decision-making to reduce patient and caregiver barriers to engage in HaH programs.

Participants expressed comfort with managing their medication and equipment during HaH. The challenges COVID-to-Home patients encountered obtaining medications from external pharmacies highlighted the need for streamlined medication processes to minimize disruptions in medication schedules. Pharmacy processes like daily provision of medications using, for example, pill packs or self-dispensing timed medication units might reduce patient and caregiver dependence on external pharmacies. Pharmacy services in a HaH program previously described ^30^ could guide the development of these processes. Caregiver availability in assisting with program components and providing emotional support is crucial for providing patient support and positive program outcomes.

Barriers to engagement included fear of the COVID-19 pandemic and caregivers’ previous experiences. Caregiving affected caregivers’ mental health with feelings of depression, loneliness, and the need for social interaction described. This finding aligns with previous reports of caregiver burden in caregivers of non-hospitalized patients, ^31,32^ with caregivers reporting anxiety, strain, and depression. ^33^ HaH programs could provide resources such as pastoral care and access to supportive individuals to alleviate caregiver distress. Patient and caregivers indicated negative HaH experiences due to strict visitation policies, limited time with loved ones, and the emotional and physical toll of prolonged hospital stays. Yet, some participants hat hospitalization provided an opportunity for rest and healing away from competing priorities at home. Future HaH programs should provide adequate mental health support and consider the potential benefits of care at home.

Some study limitations include the risk of response bias and social desirability. Asking neutral questions in a non-threatening way and using unbiased language mitigated these risks. Also, participant confidentiality was ensured during phone interviews by using a room with a closed door and with no other person present. Though telephone interviews are less costly, the absence of visual cues to guide the interviewer and inability to observe when participants are distracted are limitations. ^34^ Also, the views reported here include a very small number of those who were eligible and did not participate in HaH(n=3). Therefore, the views reported here may not represent the general views of the larger population of patients who might be eligible but did not participate in such programs.

## CONCLUSION

This study elicited patient and caregiver perceptions and organized them into a conceptual model of factors affecting patient and caregiver engagement in HaH programs. Future HaH programs should consider these factors to enhance patient and caregiver engagement and optimize implementation and clinical outcomes.

## SUPPORTING INFORMATION (APPENDIX)

### Appendix

Figure 1: Conceptual Framework for Understanding Willingness to Engage with the HaH Program

Table 1: COVID-to-Home and Hospital at Home Program

Table 2: Patient and Caregiver Interview Question Domains

Table 3: Major Qualitative Categories and Sub-categories

Table 4: Qualitative themes and representative quotes

## Data Availability

The data that support the findings of this study are available from the corresponding author upon reasonable request and with an institutional data sharing agreement in place.

## Notes

### Competing Interest Statement

The authors have declared no competing interest.

### Funding Statement

Funding for this project was provided by the Vanderbilt Institute for Clinical and Translational Research (VR55752). The Vanderbilt Institute for Clinical and Translational Research (VICTR) is funded by the National Center for Advancing Translational Sciences (NCATS) Clinical Translational Science Award (CTSA) Program, Award Number 5UL1TR002243-03. The content is solely the responsibility of the authors and does not necessarily represent the official views of the NIH.

### Author Declarations

Ethics committee/IRB of Vanderbilt University Medical Center gave ethical approval for this work

